# Effectiveness of inactivated COVID-19 vaccines against SARS-CoV-2 infections among healthcare personnel in Pakistan: a test-negative, case-control study

**DOI:** 10.1101/2023.01.09.23284342

**Authors:** Unab I. Khan, Imran Hassan, Mahnoor Niaz, Syed Iqbal Azam, Zahra Hasan, Syed Faisal Mahmood, Asad Ali

## Abstract

**Objective:** During the COVID-19 pandemic, several vaccines that were efficacious in randomized controlled trials (RCTs) were authorized for mass vaccination. In developing countries, inactivated vaccines were widely administered. While inactivated vaccines have been deemed effective in reducing disease severity, for healthcare personnel (HCPs), effectiveness against COVID-19 infections is also essential to reduce the risk to vulnerable patients and ensure a stable healthcare workforce. In addition, there are limited studies examining inactivated vaccines’ effectiveness against emerging SARS-CoV-2 variants in real-world settings. We aimed to estimate the effectiveness of inactivated vaccines (BBIBP-CorV and CoronaVac) against RT-PCR-confirmed COVID-19 infections among HCPs in the setting of emerging SARS-CoV-2 variants in Pakistan.

**Design, setting and participants:** A retrospective matched test-negative case-control analysis of existing data of HCPs at a private healthcare system in Pakistan.

**Methods:** HCPs tested between April 1 and September 30, 2021, were included. Each case was matched to two to six controls by the date of the RT-PCR test (± 7 days) to reduce bias. We compared demographics, reasons for testing, and vaccination status between cases and controls using chi-square for categorical variables and t-test for continuous-level data. The odds of getting a PCR-confirmed SARS-COV-2 infection were calculated using conditional logistic regression, after adjusting for age, gender, and work area. Vaccine effectiveness (VE) was calculated as percent VE using (1-OR)*100.

**Results:** Inactivated vaccines were ineffective against COVID-19 infections ≥ 14 days after receiving the first dose [VE: 20% (95% CI: −10, 41; p=0.162)]. The vaccines showed modest effectiveness ≥ 14 days after the second dose against COVID-19 infections [VE: 33% (95% CI: 11, 50; p=0.006)], and symptomatic COVID-19 infections [VE: 36% (95% CI: 10, 54; p=0.009)].

**Conclusions:** Inactivated vaccines show modest effectiveness against COVID-19 infections in the setting of emerging VOCs. This builds a strong case for boosters and/or additional vaccination.

## INTRODUCTION

The coronavirus disease 2019 (COVID-19) pandemic impacted global health, and as of December 9, 2022, more than 657 million COVID-19 cases and six million deaths have been reported to the WHO (1). Mass vaccination remains a cornerstone of public health interventions to counter the COVID-19 pandemic. Several vaccines have proven efficacious in phase III trials and received emergency approval for mass vaccination campaigns (2-4). As healthcare personnel (HCPs) are at a higher risk of contracting the disease and can become a source of infection to vulnerable patients (5, 6), both Centers for Disease Control (CDC) and WHO (World Health Organization) have recommended that national vaccine strategies prioritize vaccination of HCPs (7, 8).

In Pakistan, the Government-driven vaccination campaign commenced in February 2021. Vaccines with more than 50% efficacy in clinical trials were approved for mass vaccination by the health regulatory authorities, and HCPs were prioritized for vaccination (9). In the first phase, conventional inactivated vaccines BBIBP-CorV and CoronaVac were administered. Over the next few months, replication-deficient adenovector vaccines, including single-dose Ad5-nCoV (CanSino Bio) and the two-dose ChAdOx1 nCoV-19 (Oxford-AstraZeneca), became available and were administered to HCPs and the public (10).

While CoronaVac and BIBP-CorV were efficacious in clinical trials (11, 12) most were conducted before SARS-CoV-2 variants of concern (VOCs) appeared. It, therefore, became crucial to assess the effectiveness of these vaccines in real-world settings during the emergence of VOCs. This information can provide critical insights to help with policy decisions, including the need to give boosters or additional vaccination. We report the effectiveness of BBIBP-CorV and CoronaVac vaccines against RT-PCR-confirmed COVID-19 infections among HCPs, four months into the vaccination drive at a large private healthcare system in Pakistan, with an existing robust employee surveillance system. Our study was conducted in the setting of the third and fourth waves of COVID-19 in Pakistan during which the alpha, beta, gamma, and delta variants were prevalent (13, 14).

## METHODS

### Study Design

We conducted a matched, test-negative case-control study to evaluate the effectiveness of inactivated vaccines (BBIBP-CorV and CoronaVac) in reducing the odds of RT-PCR-confirmed COVID-19 infections in HCPs. Due to the rapidly changing vaccine uptake and prevalence of the disease in the community, we used the test-negative case-control design as recommended by the WHO (15). Our study population included all HCPs working at AKU who were tested for SARS-CoV-2 by RT-PCR at the AKU Hospital Clinical Laboratories between April 1 and September 30, 2021.

De-identified data was transferred from the Employee Health database to STATA v.15.0 for analysis. This study was approved by the Ethical Review Committee (ERC#: 2021-5629-18170), AKU.

### Study Population

#### Inclusion Criteria

- HCPs working at AKU and who got an RT-PCR test for SARS-CoV-2 at AKU Hospital Clinical Laboratories between April 1 and September 30, 2021, AND
- HCPs who were unvaccinated or vaccinated with one or two doses of BBIBP-CorV or CoronaVac.

#### Exclusion Criteria

- HCPs with missing vaccination data.
- HCPs who received other vaccines (Ad5-nCoV, AZD1222 (ChAdOx1), mRNA-1273, Janssen (Johnson & Johnson), Gam-COVID-Vac, BNT162b2).
- HCPs who were enrolled in COVID-19 vaccine trials.

### Study Setting

The Aga Khan University (AKU), a not-for-profit organization, runs a large healthcare system within Pakistan. Its main campus in Karachi is a 750-bed tertiary care hospital, a medical college, and a nursing school. Additionally, four secondary-care hospitals in two cities, 19 integrated medical centers, and 290 laboratory collection centers in 120 cities across Pakistan are part of the AKU healthcare system. The University employs 13960 staff, of which 80% are involved in direct health care (16).

Since the beginning of the pandemic, multiple policies were put in place to facilitate HCPs getting tested. Free assessments, testing, and treatment were offered through the Office of Employee Health in the Department of Family Medicine. In addition, time away from work due to quarantine and isolation was not counted from HCPs’ annual leaves. For each employee who tested positive, detailed contact tracing was performed. With these employee-friendly policies, HCPs have utilized the Office of Employee Health; to date, 30,000 tests have been conducted.

HCPs were tested if: 1) they had symptoms consistent with COVID-19; 2) had a high-risk exposure, as defined by the CDC criteria (17), to a person infected with COVID-19 either in the community or at the workplace; or 3) were part of an outbreak investigation in a specific part of the University. All testing was performed by SARS-CoV-2 polymerase chain reaction on a nasal specimen using the Cobas® 6800 Roche assay (18). The samples were collected at the HCPs’ workplace, and all testing was done at the AKU Hospital Clinical Laboratories, accredited by the College of American Pathologists, USA (16).

Office of Employee Health maintained a password-protected database for COVID-19-related data that is separate from HCPs’ medical records. Once vaccination began, HCPs also provided dates of vaccine administration and the type of vaccine. Vaccine information was confirmed by verifying with the national database using the SMS-based system developed by the Government of Pakistan (10). Vaccinations were available without any priority policy to all AKU HCPs, regardless of age, comorbidities, area of work, previous SARS-CoV-2 infections, etc.

### Definitions

#### Healthcare Personnel

All employees working within the healthcare system with the potential of direct and indirect exposure to patients or infectious material were considered HCPs (19).

#### Case

HCPs who had a positive SARS-CoV-2 RT-PCR test during the study period and had an absence of a positive test result in the preceding 90-day period.

#### Control

HCPs with a negative SARS-CoV-2 RT-PCR test result during the study period and the absence of a positive test result in the preceding 90-day and the subsequent 14-day period.

#### Vaccination Status

Using the WHO definition (20), we defined vaccination status at the time of testing as:

- Unvaccinated: If no dose of any vaccine was received.
- Single dose received: If only the first dose of BBIBP-CorV or CoronaVac vaccine was received. This group was further divided into two subgroups: a) 0–13 days since receiving the first dose; b) ≥ 14 days since receiving the first dose of a two-dose vaccine.
- Two doses received: If both doses of BBIBP-CorV or CoronaVac vaccine were received. This group was also further divided into two: a) 0–13 days since receiving the final dose; b) ≥ 14 days since receiving the final dose.

#### COVID-19 Infection

All HCPs who tested positive for SARS-CoV-2 by RT-PCR, regardless of the presence or absence of symptoms.

#### Symptomatic COVID-19

All HCPs who tested positive for SARS-CoV-2 by RT-PCR and had one or more COVID-19-related symptoms in 0-10 days before the RT-PCR test for SARS-CoV-2.

#### Work Area

HCPs not working in direct clinical care (e.g. laboratory personnel, housekeeping staff, food providers, and administrative staff) were classified as “non-clinical”; those providing direct clinical care in areas not designated for patients with COVID were classified as “clinical non-COVID”; whereas HCPs working in designated areas for COVID-suspect or COVID-confirmed patients were categorized as “clinical COVID.”

### Statistical analysis

Data on demographics, results of the SARS-CoV2 RT-PCR test, vaccination status and dates, reason for testing, work area, and previous RT-PCR tests (within the preceding 90-day period) were retrieved from the Office of Employee Health’s database for statistical analysis. Vaccination status was allocated based on pre-defined definitions stated above.

As the study was during the surge, with percent positivity changing weekly, we matched cases and controls by the date of the RT-PCR test (± 7 days) to reduce bias. Each case was matched to a minimum of two and a maximum of six controls.

We compared demographics, reason for testing, work area and vaccination status between cases and controls using chi-square for categorical variables and t-test for normally distributed continuous-level data. Using unvaccinated individuals as a reference, we used conditional logistic regression to estimate the odds of having RT-PCR-confirmed COVID-19 infections in HCPs vaccinated with a single dose and both doses of the two-dose inactivated vaccines. We calculated unadjusted and adjusted odds ratios, accounting for covariates that included age, sex (using females as a reference), and work area (using non-clinical as a reference). Vaccine effectiveness was calculated as Percent VE using (1-OR)*100. Additionally, we performed a subgroup analysis to assess the effectiveness of vaccines against symptomatic COVID-19 infection.

Based on Hitchings et al study (21), we plotted the weekly positivity rate and cumulative vaccine coverage from the time vaccines were introduced (January 2021) to the end of the study period (end of delta surge in September 2021) to understand the impact of vaccines on COVID-19 infections in the healthcare system.

### Patient and public involvement

Patient and the public were not involved in the conduct of this study.

## RESULTS

Between April 1 and September 30, 2021, 4599 HCPs were tested for SARS-COV-2 via RT-PCR. Figure 1 shows the process for the selection of HCPs in analysis. After exclusion, 4074 HCPs remained, of whom 1037 tested positive and were classified as cases, and 3037 tested negative and were classified as controls. Of these, 3095 (959 cases and 2136 controls) were tested for symptoms. Cases were matched to controls by the date of the RT-PCR test (± 7 days). Each case was matched to two to six controls. No HCPs had to be excluded due to non-matching.

**Figure 1.**
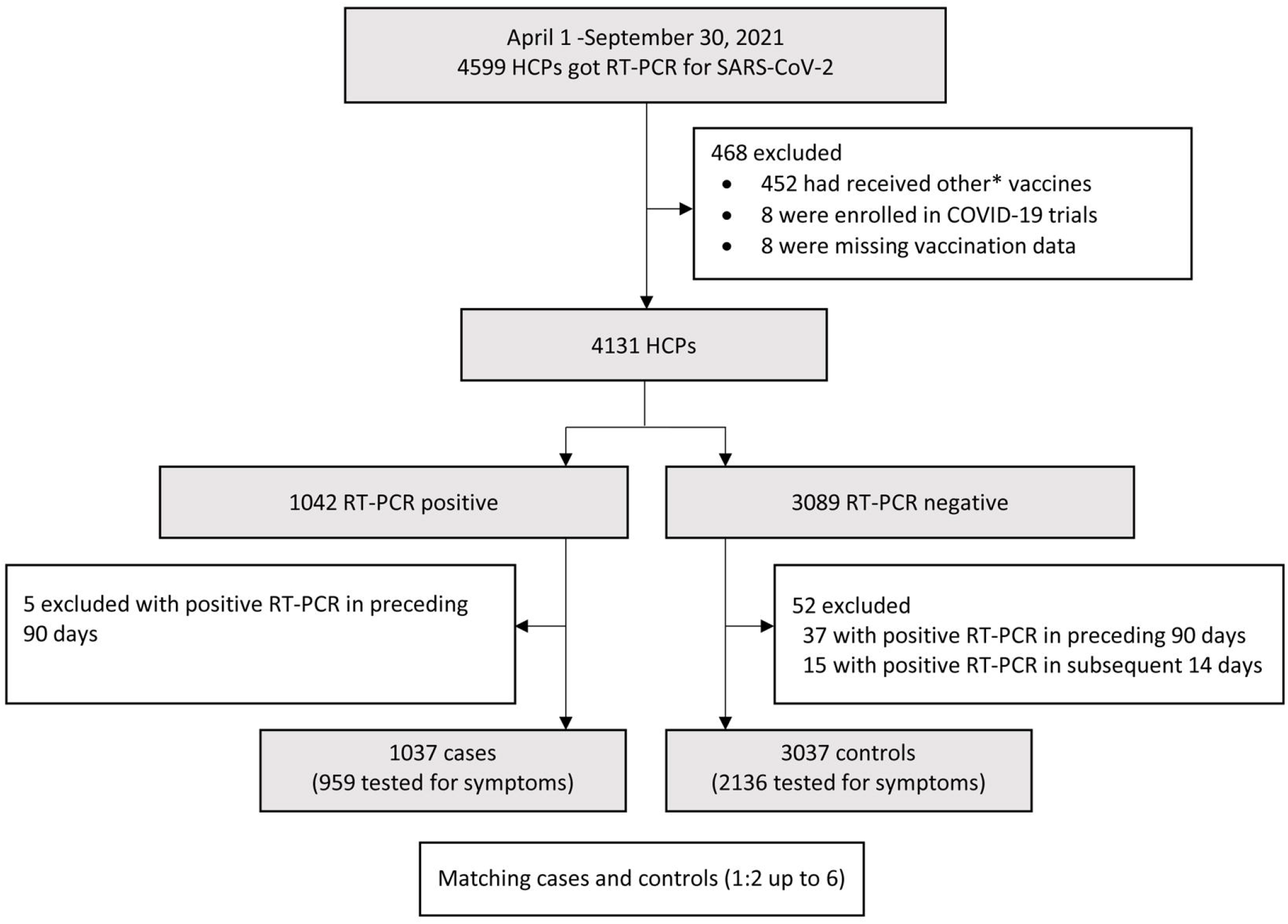
Process for the selection of HCPs. *Other vaccines: Ad5-nCoV (n=342), AZD1222 (ChAdOx1) (n=60), mRNA-1273 (n=20), Janssen (n=1), Gam-COVID-Vac (n=18), BNT162b2 (n=11).

Table 1 shows the characteristics of all cases and controls. Compared to controls, cases were older (35.3 ± 9.7 vs. 33.1 ± 8.6; p-value < 0.001). 73.9% of cases and 74.8% of controls were administered the BBIBP-CorV vaccine. 61.4% of cases and 60.3% of controls had received the second dose at least 14 days before the test. Among HCPs that had received both doses of vaccine before the RT-PCR test, there was no significant difference in mean duration between the second dose and test between cases and controls (94.8 ± 41.6 vs. 91.7 ± 45.1 days; p value=0.123).

**Table 1.**
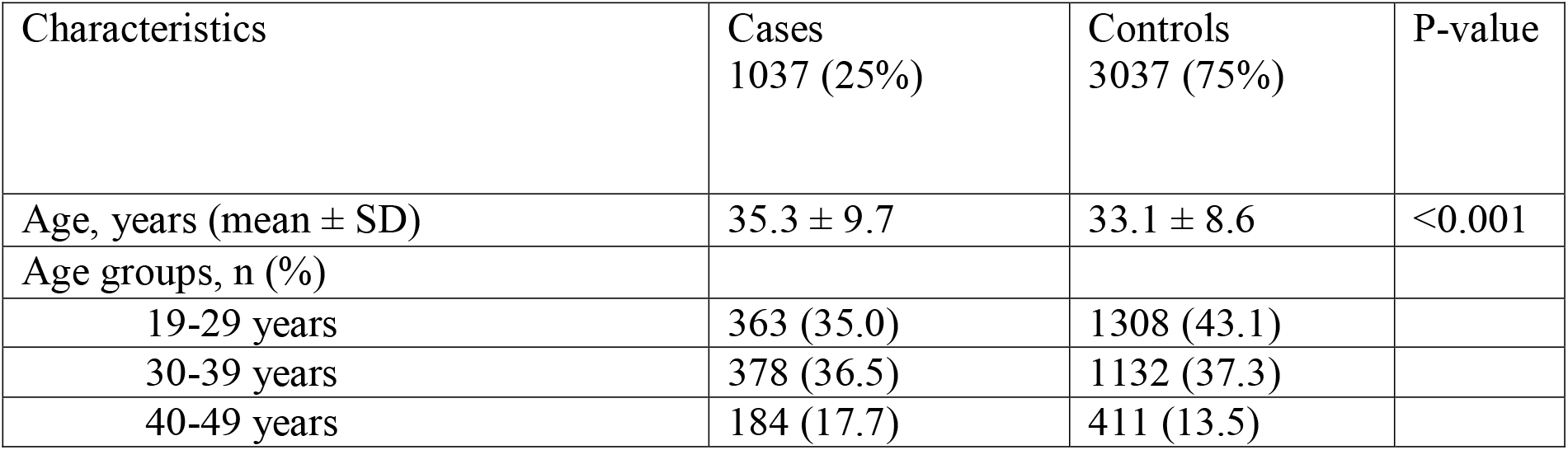
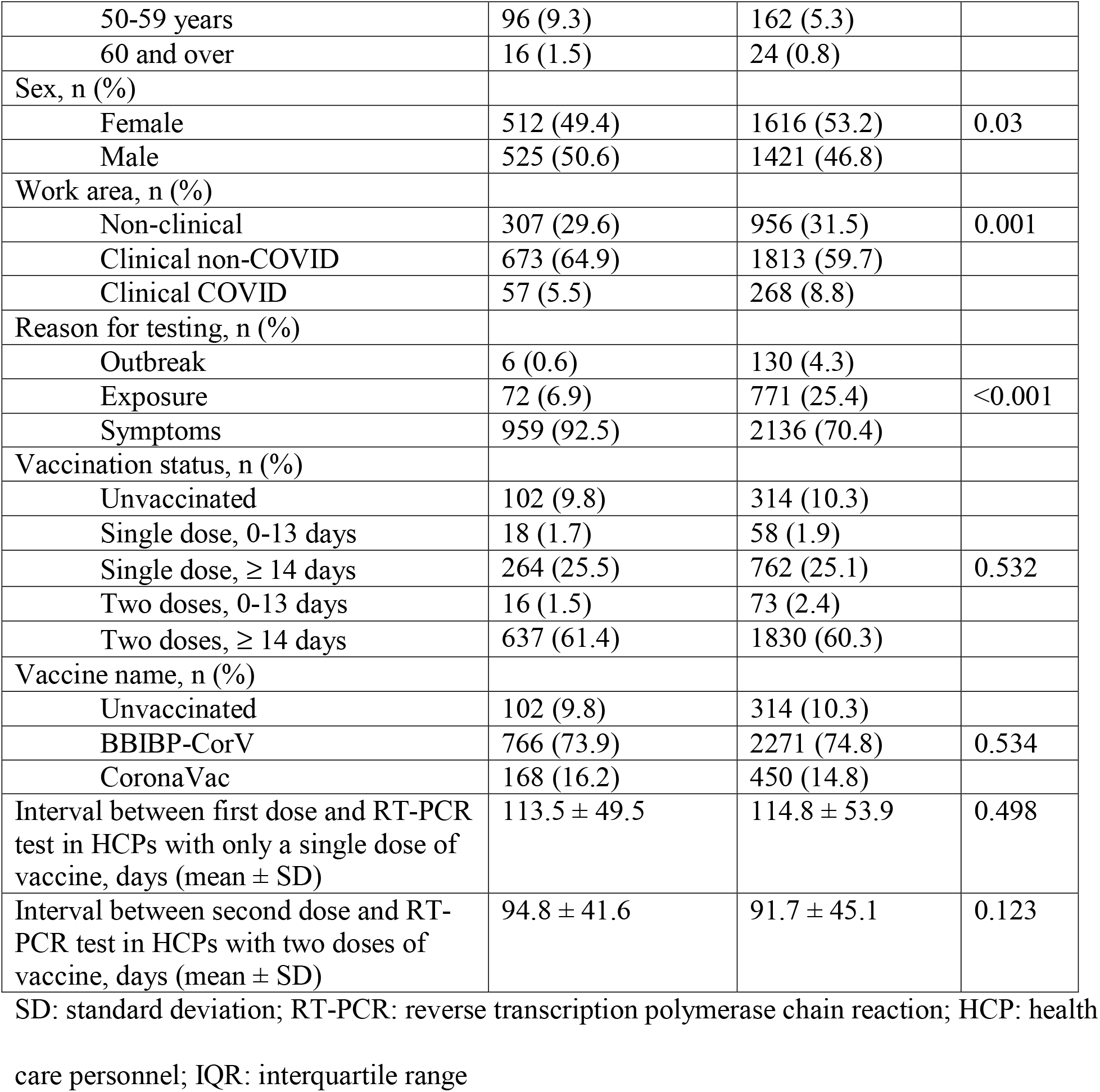
Characteristics of all cases and controls (N=4074)

Table S1 describes the characteristics of symptomatic cases and controls (3095/4074 (76.0%). There was no significant difference in the mean interval between the onset of symptoms and the RT-PCR test between cases and controls (2.1 ± 2.0 vs. 1.9 ± 2.3 days; p=0.017).

Table 2 shows the odds of contracting COVID-19 infection by the time since vaccination. After adjusting for age, sex, and work area, we found that a single dose of the two-dose vaccines was ineffective within the first 13 days [VE: 30% (95% CI: −26, 62; p=0.234)] or even after 13 days had elapsed [VE: 20% (95% CI: −10, 41; p=0.162)]. This shows that one dose is not enough to prevent COVID-19 infections. While the vaccines were effective during the period < 14 days after the second dose [VE: 44% (95% CI: −2, 69; p=0.055)], the results were not significant. The vaccines were most effective ≥ 14 days after the second dose against a COVID-19 infection [VE: 33% (95% CI: 11, 50; p=0.006)].

**Table 2.**
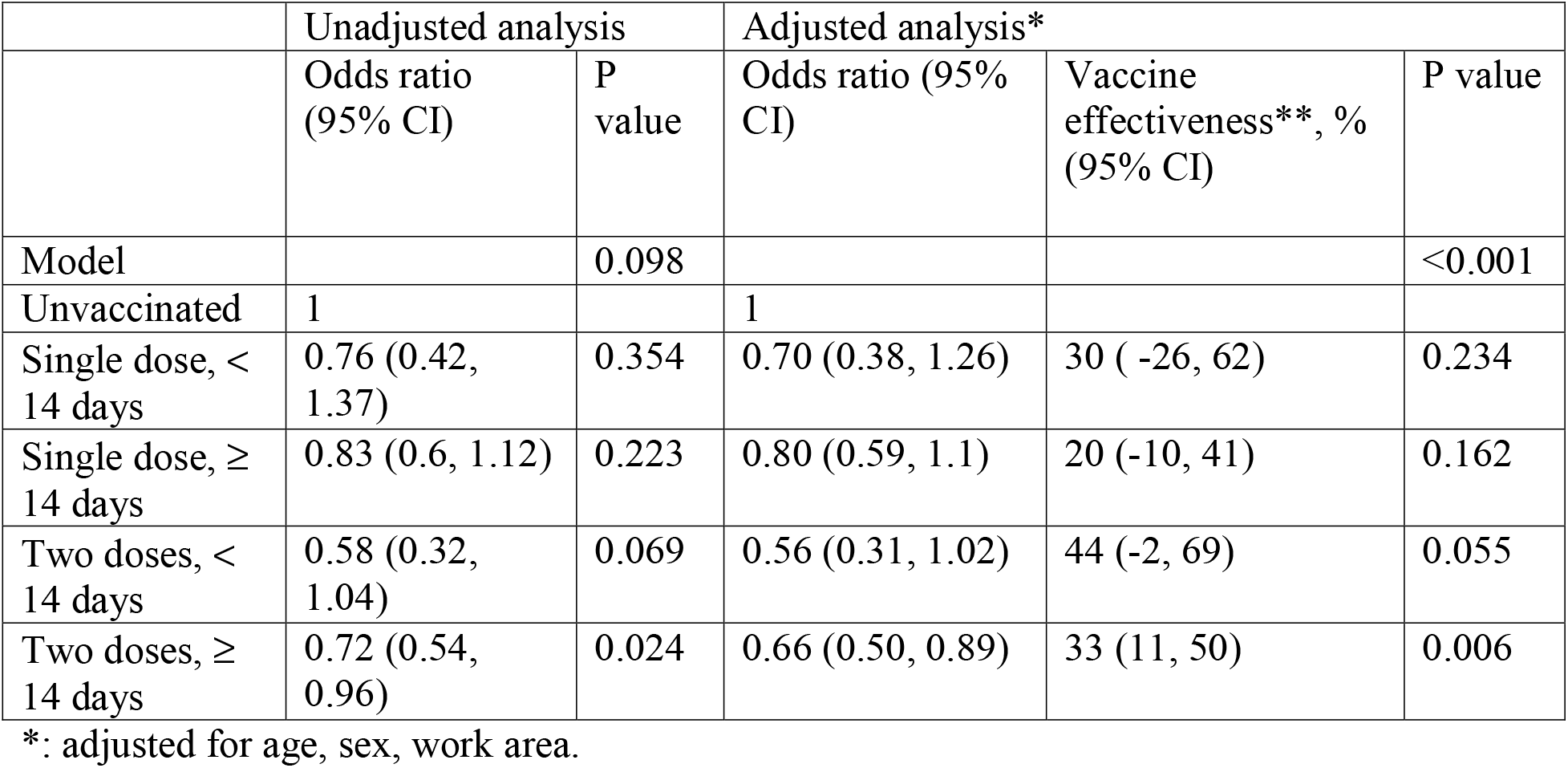
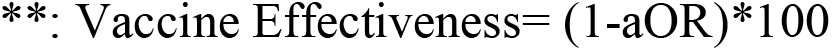
Effectiveness of inactivated vaccines against all COVID-19 infections (symptomatic and asymptomatic) (matched case-control)

|  | Unadjusted analysis |  | Adjusted analysis* |  |  |
| --- | --- | --- | --- | --- | --- |
|  | Odds ratio (95% CI) | P value | Odds ratio (95% CI) | Vaccine effectiveness**, % (95% CI) | P value |
| Model |  | 0.098 |  |  | <0.001 |
| Unvaccinated | 1 |  | 1 |  |  |
| Single dose, < 14 days | 0.76 (0.42, 1.37) | 0.354 | 0.70 (0.38, 1.26) | 30 ( -26, 62) | 0.234 |
| Single dose, ≥ 14 days | 0.83 (0.6, 1.12) | 0.223 | 0.80 (0.59, 1.1) | 20 (-10, 41) | 0.162 |
| Two doses, < 14 days | 0.58 (0.32, 1.04) | 0.069 | 0.56 (0.31, 1.02) | 44 (-2, 69) | 0.055 |
| Two doses, ≥ 14 days | 0.72 (0.54, 0.96) | 0.024 | 0.66 (0.50, 0.89) | 33 (11, 50) | 0.006 |

\*: adjusted for age, sex, work area.
\*\*: Vaccine Effectiveness= (1-aOR)\*100

Table 3 shows the odds of contracting symptomatic COVID-19 infection by the time since vaccination. After adjusting for age, sex, and work area, we found that the first dose was ineffective against a symptomatic COVID-19 infection. The vaccines were most effective ≥ 14 days after the second dose against a symptomatic COVID-19 infection [VE: 36% (95% CI: 10, 54; p=0.009)].

**Table 3.** Effectiveness of inactivated vaccines against symptomatic COVID-19 infections (matched case-control)

|  | Unadjusted analysis |  | Adjusted analysis* |  |  |
| --- | --- | --- | --- | --- | --- |
|  | Odds ratio<br>(95% CI) | P<br>value | Odds ratio (95%<br>CI) | Vaccine<br>effectiveness**, %<br>(95% CI) | P value |
| Model |  | 0.208 |  |  | <0.001 |
| Unvaccinated | 1 |  | 1 |  |  |
| Single dose, <<br>14 days | 0.82 (0.42,<br>1.57) | 0.546 | 0.71 (0.36, 1.38) | 29 (-38, 64) | 0.295 |
| Single dose, ≥<br>14 days | 0.76 (0.53,<br>1.08) | 0.125 | 0.75 (0.52, 1.07) | 25 (-7, 48) | 0.101 |
| Two doses, <<br>14 days | 0.60 (0.31,<br>1.15) | 0.122 | 0.60 (0.31, 1.16) | 40 (-16, 69) | 0.131 |
| Two doses, ≥<br>14 days | 0.69 (0.5, 0.96) | 0.027 | 0.64 (0.46, 0.90) | 36 (10, 54) | 0.009 |

\*: adjusted for age, sex, work area.
\*\*: Vaccine Effectiveness= (1-aOR)\*100

Figure 2 displays the weekly RT-PCR tests conducted and the cumulative coverage of the first and second dose of inactivated vaccines among HCPs at our institution. We see that despite 75% coverage with 2-doses of inactivated vaccines, we observed an increase in positivity rates that correlated with the delta surge in Pakistan. This also shows the modest effectiveness of inactivated vaccines against VOCs.

**Figure 2.**
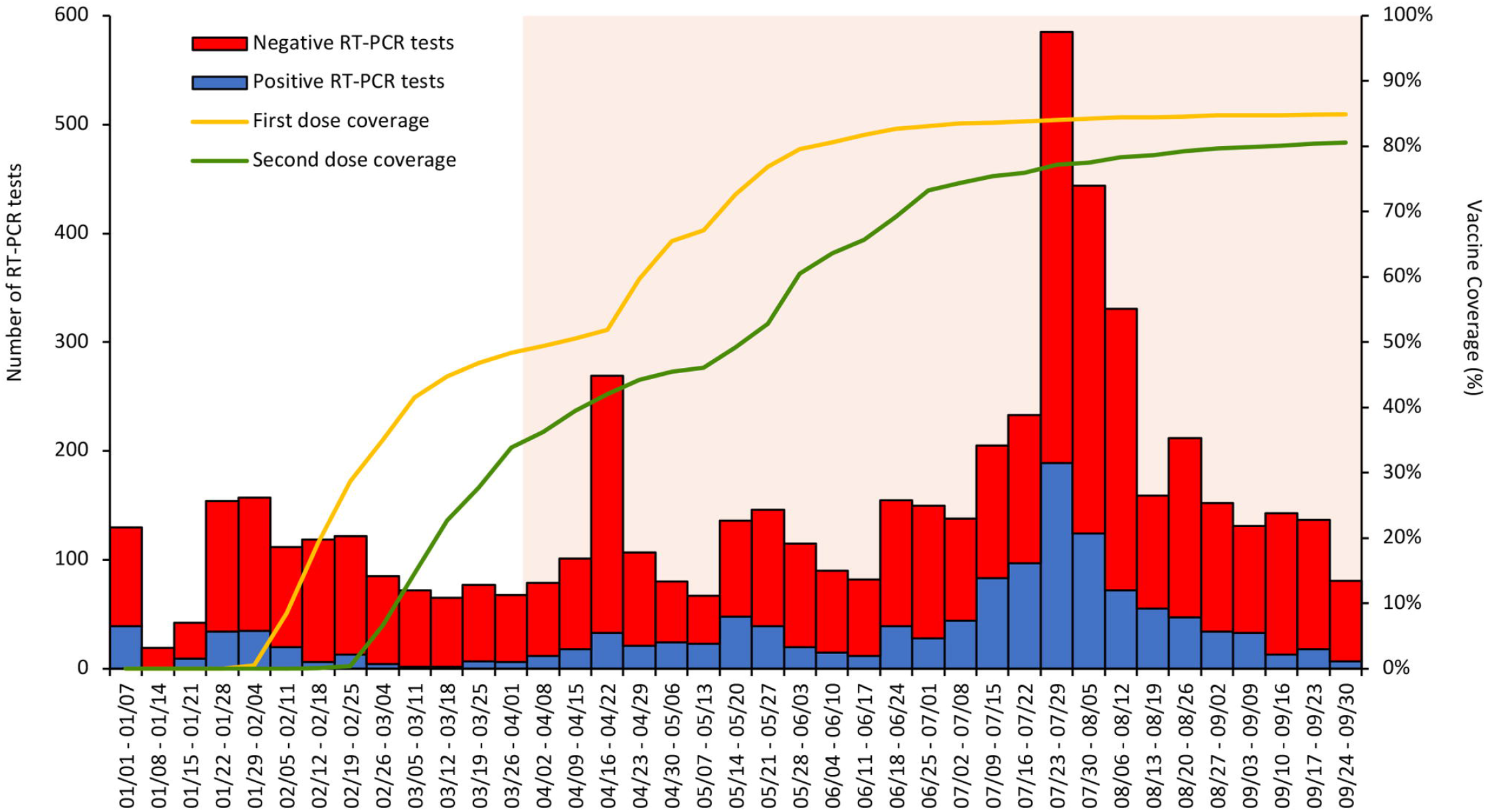
Weekly number of RT-PCR tests conducted and cumulative coverage of inactivated vaccines (BBIBP-CorV and CoronaVac) among HCPs at our institution from January 1-September 30, 2021. Negative and positive RT-PCR tests are represented as red and blue bars, respectively. Cumulative coverage of first and second dose of inactivated vaccines is shown by the yellow and green lines, respectively. Our study period, April 1-September 30, 2021, is highlighted in pink.

## DISCUSSION

Our study among HCPs found that BBIBP-CorV and CoronaVac were not as effective as seen in the efficacy trials (11, 12) in preventing COVID-19 infections, even 14 days after the second dose of the two-dose vaccines. Vaccines did not provide significant protection against infection after the first dose and within the first two weeks of the second dose. During our study period, the predominant variants in Pakistan switched from alpha, beta and gamma in April to June (13, 14) to the highly transmissible delta constituting up to 99% of the total genomes sequenced from July to September 2021 (22).

Existing evidence on the effectiveness of inactivated vaccines against infections is inconsistent. Our results agree with a test-negative case-control study conducted in Brazil among healthcare workers during the gamma variant epidemic, which reported modest effectiveness of two doses of CoronaVac, with adjusted effectiveness of 37.1% against symptomatic COVID-19 infections and 37.9% against all COVID-19 infections (21). Similarly, moderate effectiveness against COVID-19 infections was estimated for inactivated vaccines during the delta variant epidemic in China (23, 24). In contrast, the effectiveness estimated in our study is lower than that of CoronaVac reported among healthcare workers in Turkey (65%) (25).

Several possibilities exist for the lower-than-expected effectiveness of inactivated vaccines in our study. Neutralizing antibody levels correlate with protection against COVID-19 infections (26, 27), and inactivated vaccines produce inferior antibody responses compared to mRNA vaccines (9, 28-30). Moreover, the weakened neutralization potency of inactivated vaccines against the dominant VOCs circulating during our study period, especially the vaccine-resistant delta variant, may have contributed to the limited effectiveness (30, 31). Indeed, studies conducted during the delta variant epidemic have demonstrated only modest effectiveness of two doses of BBIBP-CorV and CoronaVac against infections (23, 32).

The mean interval between the second dose of vaccine and COVID-19 infection was more than 90 days in our study. Studies have demonstrated a decline in neutralizing antibody titers three months after vaccinations with the two-dose BBIBP-CorV and CoronaVac (30, 33, 34). It is possible that there was a significant waning of neutralizing antibody responses in our study population. Considering that inactivated vaccines were the most widely used vaccines in low-middle-income countries (LMICs), this finding supports the provision of homologous or heterologous booster doses among recipients of these vaccines.

Our study has limitations. Our sample size did not allow us to match more than one variable. However, we did adjust for age, sex, and work area in the logistic model. Additionally, most HCPs in our setting did not get the severe disease or require hospitalization during the study period. Therefore, we cannot comment on vaccines’ effectiveness against severe disease. It is, however, essential to note that in healthcare settings, infected HCPs can risk spreading the infection to vulnerable patients. Thus, it is vital to examine not only from the perspective of the infected person but its impact on the system at large. As samples from HCPs in our study population did not undergo genomic sequencing, we could not directly ascertain effectiveness against the circulating VOCs. Finally, our study was conducted on HCPs and the results may not be generalizable to the population outside of healthcare settings.

Despite these limitations, our study has clear strengths. Our findings are from a large, well-defined group of HCPs with consistent testing parameters that did not change during the study period. We used RT-PCR which is sensitive to detecting SARS-CoV-2 infections (18). Moreover, the vaccine status was highly accurate as it was validated through the national vaccine registry. Using a test-negative design, we mitigated the risk of bias/confounding associated with healthcare access and seeking behavior. Finally, our study was conducted among HCPs whose well-being is imperative to continue to offer uninterrupted, quality care to patients in a pandemic.

Our study was conducted in an LMIC with a high population density and limited resources. Thus, examining the effectiveness of vaccines in these settings is essential to prevent public health crises. Pakistan and other developing economies with weak healthcare systems need effective vaccines to meet the unprecedented challenge of dealing with the morbidity and mortality associated with COVID-19. Our study adds new estimates to the effectiveness of these vaccines in HCPs. It may guide future policies in Pakistan and other LMICs that mainly administered inactivated vaccines in their mass vaccination campaigns. By finding complete vaccination with CoronaVac and BBIBP-CorV vaccines to be only modestly effective against COVID-19 infections, our study strengthens the case for boosters or additional vaccination among recipients of these vaccines. Additionally, with new variants being identified regularly, it is crucial to prioritize conducting studies at different times and in various settings to continuously examine the effectiveness of these vaccines and boosters against the prevailing variants in real-world settings.

## CONCLUSIONS

Absence of strong effectiveness of inactivated vaccines against COVID-19 infections among HCPs, especially during the spread of VOCs in our setting is concerning and builds a case for boosters and/or additional vaccination. There is a need for further studies to continuously assess effectiveness in the setting of emerging variants to guide policies regarding boosters to ensure adequate protectiveness.

## Data Availability

As this is employee-related data, we do not have permission to make it publicly accessible. However, data are available from the corresponding author after approval from the University ethics review committee on reasonable request.

## Author Contributions

Conception and design: UIK, IH, AA

Analysis and Interpretation: UIK, IH, IA, MN

Drafting and Revising manuscript: MN, ZH, SFM, AA, IH, UIK

Final approval of manuscript: UIK

## Acknowledgments

We acknowledge the support of Professor Adil Haider, Dean AKU-Medical College and Professor Shahid Shafi, previous CEO of Aga Khan University Hospital, Pakistan. In addition, we thank the Employee Health team Shehreen Somani RN, Sania Nawaz RN, Noorjehan Momin RN, Dr. Asif Hakim, and the entire Family Medicine team for providing care to employees.

## Ethics Statements

### Participants consent to participate

Deidentified data was used for analysis and consent was not required.

### Ethics Approval

This study was approved by the Ethical Review Committee (ERC#: 2021-5629-18170), Aga Khan University.

### Competing interests

All authors declare no conflicts of interest, support or financial relationship with any organization or other activities with any influence on the submitted work.

### Data availability

As this is employee-related data, we do not have permission to make it publicly accessible. However, data are available from the corresponding author after approval from the University’s ethics review committee on reasonable request.

### Funding

This research received no specific grant from any funding agency in the public, commercial or not-for-profit sectors.

## Notes

### Competing Interest Statement

The authors have declared no competing interest.

### Author Declarations

Ethical Review Committee of Aga Khan University exempted this study because anonymised data was used for analysis.

