## Supplementary material for "Effectiveness of inactivated COVID-19 vaccines against SARS-CoV-2 infections among healthcare personnel in Pakistan: a test-negative, case-control study": Table S1

Characteristics of symptomatic cases and controls (N= 3095)

| Characteristics | Cases  (n= 959) | Controls  (n= 2136) | P value |
| --- | --- | --- | --- |
| Age, years (mean ± SD) | 35.0 ± 9.5 | 32.5 ± 8.3 | <0.001 |
| Age groups, n (%) |  |  |  |
| 19-29 years | 339 (35.4) | 973 (45.6) |  |
| 30-39 years | 357 (37.2) | 790 (37.0) |  |
| 40-49 years | 166 (17.3) | 258 (12.1) |  |
| 50-59 years | 85 (8.9) | 99 (4.6) |  |
| 60 years and over | 12 (1.2) | 16 (0.8) |  |
| Sex, n (%) |  |  |  |
| Female | 469 (48.9) | 1115 (52.2) | 0.09 |
| Male | 490 (51.1) | 1021 (47.8) |  |
| Work area, n (%) |  |  |  |
| Non-clinical | 273 (28.5) | 535 (25.1) |  |
| Clinical non-COVID | 54 (5.6) | 227 (10.6) | <0.001 |
| Clinical COVID | 632 (65.9) | 1374 (64.3) |  |
| Vaccination status, n (%) |  |  |  |
| Unvaccinated | 93 (9.7) | 190 (8.9) |  |
| Single dose, 0-13 days | 17 (1.8) | 42 (2.0) |  |
| Single dose, ≥ 14 days | 249 (26.0) | 581 (27.2) | 0.565 |
| Two doses, 0-13 days | 16 (1.7) | 52 (2.4) |  |
| Two doses, ≥ 14 days | 584 (60.9) | 1271 (59.5) |  |
| Vaccine name, n (%) |  |  |  |
| Unvaccinated | 93 (9.7) | 190 (8.9) |  |
| BBIBP-CorV | 709 (73.9) | 1610 (75.4) |  |
| CoronaVac | 157 (16.4) | 336 (15.7) | 0.663 |
| Interval between first dose and RT-PCR test in HCPs with only a single dose of vaccine (days, mean (SD)) | 113.8 ± 49.7 | 115.5 ± 55.0 | 0.446 |
| Interval between second dose and RT-PCR test in HCPs with two doses of vaccine, (days, mean (SD)) | 95.0 ± 41.5 | 91.4 ± 45.1 | 0.094 |
| Interval between symptom onset and RT-PCR test, days (mean ± SD) | 2.1 ± 2.0 | 1.9 ± 2.3 | 0.017 |

SD: standard deviation; RT-PCR: reverse transcription polymerase chain reaction; HCP: health care personnel; IQR: interquartile range
